# Conversational AI facilitates mental health assessments and is associated with improved recovery rates

**DOI:** 10.1101/2022.11.03.22281887

**Authors:** Max Rollwage, Keno Juchems, Johanna Habicht, Ben Carrington, Tobias Hauser, Ross Harper

## Abstract

Mental health services across the globe are overburdened due to increased patient need for psychological therapies and a shortage of qualified mental health practitioners. This is unlikely to change in the short-to-medium term. Digital support is urgently needed to facilitate access to mental healthcare whilst creating efficiencies in service delivery. In this paper, we evaluate the use of a conversational artificial intelligence (AI) solution (*Limbic Access*) to assist both patients and mental health practitioners around referral, triage, and clinical assessment of mild-to-moderate adult mental illness. Assessing this solution in the context of England’s NHS Improving Access to Psychological Therapies (IAPT) services, we demonstrate that deploying such an AI solution is associated with improved recovery rates. We find that those IAPT services that introduced the conversational AI solution improved their recovery rates, while comparable IAPT services across the country reported deteriorating recovery rates during the same time period. Further, we provide an economic analysis indicating that the usage of this AI tool can be highly cost-effective relative to other methods of improving recovery rates. Together, these results highlight the potential of AI solutions to support mental health services in the delivery of quality care in the context of worsening workforce supply and system overburdening.

**Author summary:** In this paper, we evaluate the use of a conversational artificial intelligence solution (Limbic Access) to assist both patients and mental health practitioners around referral, triage, and clinical assessment of mild-to-moderate adult mental illness. Assessing this solution in the context of England’s NHS mental health services, we demonstrate that deploying such an AI solution is associated with improved recovery rates. We find that those services that introduced the conversational AI solution improved their recovery rates, while comparable mental health services across the country reported declining recovery rates during the same time period. Further, we provide an economic analysis indicating that the usage of this AI tool can be highly cost-effective relative to other methods of improving recovery rates. Together, these results highlight the potential of AI solutions to support mental health services in the delivery of quality care in the context of reduced workforce supply and an overburdened system.

## 1 Introduction

Mental illness is the largest cause of disability in western countries [Nochaiwong et al., 2021] and the COVID-19 pandemic has further fuelled this crisis [Busetta et al., 2021, Murch et al., 2021, Thome et al., 2021, Ornell et al., 2021, Marques et al., 2020, Loosen et al., 2021]. However, a lack of funding across many mental health services combined with a shortage of trained mental health practitioners has resulted in a severe supply-demand imbalance for psychological therapies. This has negative and wide-ranging consequences for those seeking help, from impoverished patient experience to worse treatment outcomes [Scott, 2018b].

A key adverse outcome from system overburdening is long waiting times between the point of patient referral and subsequent clinical assessment and treatment. For instance, in England’s National Health Service (NHS) in 2020, 30.8 % of referrals to IAPT services dropped out of the service whilst on the waiting list, before starting any treatment. 12.6 % of patients waited more than 6 weeks for an initial assessment (NHS Digital, 2020), and a further 50 % of patients experienced “hidden waits” of over 28 days between assessment and their first treatment session. This is in stark contrast to the National Institute for Health and Care Excellence (NICE) treatment guidelines that highlight a swift treatment as a key to recovery [Larsson et al., 2022]. Unfortunately, due to funding challenges and a shortage of qualified staff, workforce improvements are not to be expected in the near future [Adams et al., 2021].

To mitigate the supply-demand imbalance in mental healthcare, digital and AI solutions have been put forward as means to reduce the burden on staff and to facilitate assessment and treatment [Jayaraajan et al., 2022, Rudd and Beidas, 2020, Koutsouleris et al., 2022, Hauser et al., 2022]. Of those, AI solutions that need little-to-no input from mental health professionals are particularly suited to address the workforce crisis (in contrast to online therapy, for example, which remains labour intensive) [D’Alfonso, 2020, Ćosić et al., 2020]. Indeed, well-designed AI has the potential to augment and support human practitioners. For instance, freeing up service staff from tasks which require less clinical domain knowledge will allow them to dedicate more of their time to tasks that critically rely on clinical expertise, such as delivering therapy. AI solutions may therefore not only improve the capacity of mental healthcare services but may even improve the quality of care provided by each therapist.

An important starting point for AI solutions in mental healthcare is the referral and clinical assessment process, i.e. patient intake and initial triage of a prospective patient into the service. In most psychological therapy care pathways, the referral is a structured process that follows standardised protocols. During this process, clinical (and demographic) information may be gathered to inform a decision about the prospective patient’s suitability and to form an initial description of the patient’s presenting problem. While in practice staff intensive, this initial data collection – often standardised self-reports and questionnaires – theoretically requires minimal human oversight and is thus an ideal target for automation. In many services, this process is still conducted by clinical staff in and it is estimated that IAPT services spend up to 25 % of their budget on clinical assessments alone [Scott, 2018a]. Automation of these early stages of the care pathway represents a viable opportunity to release valuable clinical resources and improve patient treatment.

Conversational AI solutions in mental healthcare have garnered increased attention in recent years [Car et al., 2020] as they may benefit patients, clinicians and mental health services. The patient can directly benefit from harmonised data collection, through the acceleration of the intake process and through providing their clinician with a comprehensive and standardised overview of therapeutically meaningful information (improving efficiency of the practitioner-led clinical assessment and allowing more time for building therapeutic alliance and managing expectations). Indirect patient benefits may further be realised through increased efficiency of the overall service, freeing up resources for treatment. Finally, patients may benefit from improved experience of care, such as feeling less stigma around referring into mental health treatment via interaction with a non-judgemental AI rather than another human, or indeed the ability to access care at all times of the day (i.e. out of office hours). All these effects can be expected to directly or indirectly impact patient recovery rates.

Previous research into the efficiencies and effectiveness of conversational AI solutions in healthcare is limited [Wilson et al., 2022]. The majority of studies investigating chatbots in a healthcare setting often only rely on brief follow-up periods [Vaidyam et al., 2019], trials in non-clinical settings [Fitzpatrick et al., 2017], or use differing definitions of “recovery” [Meadows et al., 2020]. Thus, in this study, we examine the efficiency and effectiveness of a specific AI-enabled self-referral tool within a clinical setting, embedded as a conversational AI chatbot (*Limbic Access*). This AI-enabled self-referral tool is already embedded across multiple NHS IAPT services in the UK and therefore permits evaluation of this technology in a real-world setting [Koutsouleris et al., 2022]. Moreover, IAPT services represent a unified and structured system for access and provision of primary mental healthcare for the general adult population and thus lend themselves ideally to testing digital solutions in a controlled fashion at a large scale. This makes it an ideal test bed for evaluating digital solutions in mental healthcare more widely.

Here, we provide evidence in an unprecedented large sample (58,475 patients) of data from a real-world setting (i.e. mental health patients entering treatment) that an AI-enabled self-referral tool increases clinical efficacy by improving recovery rates compared to services without AI-enabled solutions. Moreover, an economic analysis reveals that the use of such an AI-enabled solution may be materially more economically viable than alternative solutions.

## 2 Materials and Methods

### 2.1 AI-enabled self-referral

We evaluate the effects of a novel AI-enabled self-referral tool (*Limbic Access*) which is a part of standard care in several IAPT services around the UK. This self-referral tool is a conversational AI solution which collects all relevant information required from the patient in order to refer to the IAPT service (e.g. age and location). Moreover, the self-referral tool collects further clinically relevant information, such as the Patient Health Questionnaire-9 (PHQ-9) [Kroenke et al., 2001] and Generalised Anxiety Disorder Assessment (GAD-7) [Spitzer et al., 2006], which are attached to the referral. This additional information enables the clinician to prepare for the clinical assessment and to have more context about the specific mental health issues experienced by the patient. See Figure 1B for an illustration of the referral flow.

**Figure 1:**
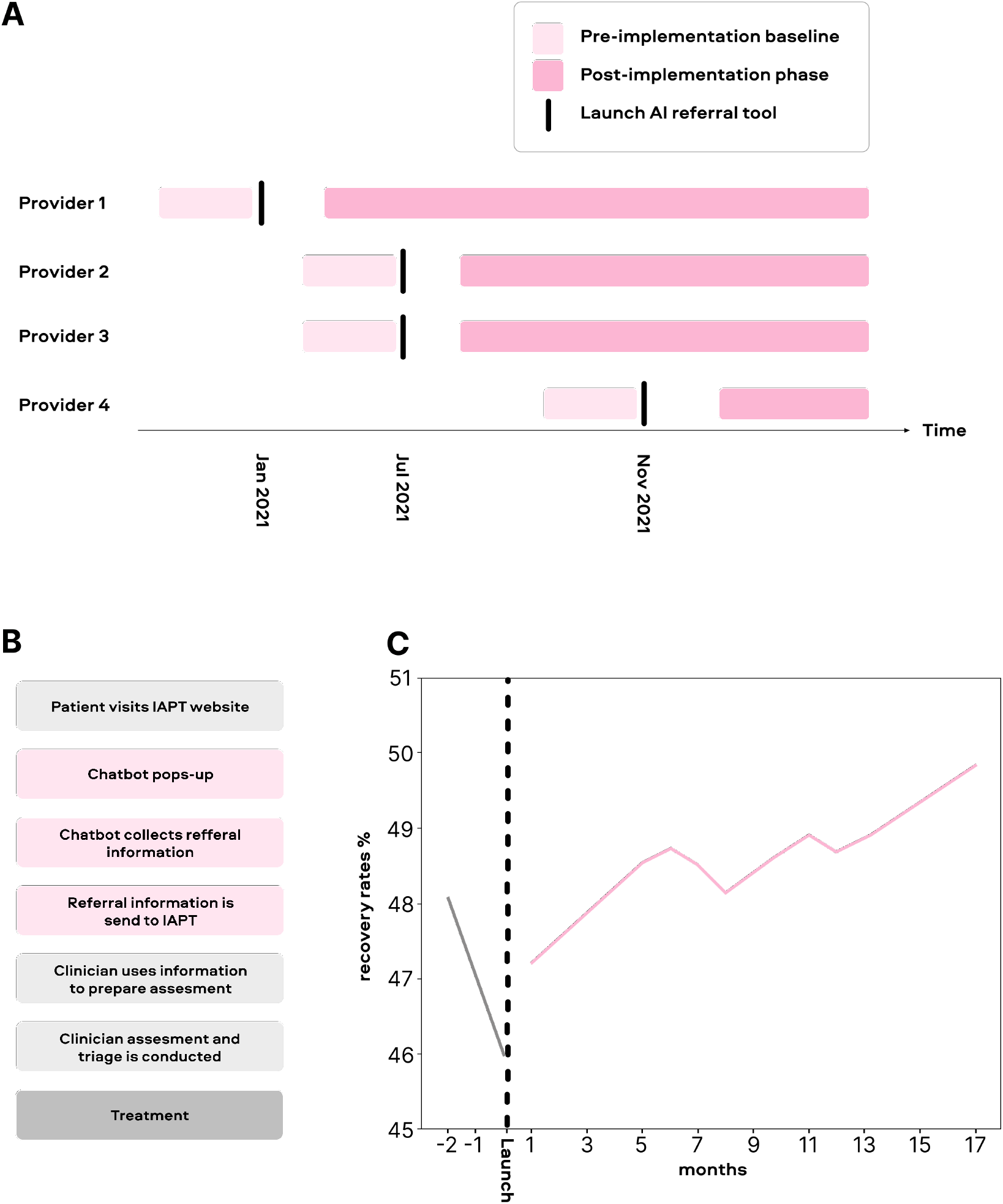
A) Study set-up showing the time of launch for the AI-enabled self-referral tool in the different IAPT services. Note that the services launched the tool at different time points. The analysis is locked to the time of implementation. This means the pre- and post-implementation period contains different time periods for different services. This rules out simple confounding effects such as external effect co-occurring with the launch of the AI-enabled self-referral tool. B) Referral workflow, showing the patient journey from visiting the IAPT website to entering treatment. C) Recovery rates before and after implementation of the AI e-referral tool. The recovery rates are averaged over all IAPT services using the AI-enabled self-referral tool. Time-locked to the time of their implementation of the 4 AI tool and recovery rates has been smoothed over time (using a Savitzky Golay filter).

### 2.2 Dataset

The data was derived from the public NHS Digital UK IAPT database, where each IAPT service reports statistics regarding their treatment outcomes and service metrics to allow for a public evaluation of service performance. This dataset has the advantage that data is available for all IAPT services of interest. As a result, no bias can occur based on missing data (e.g. services not being able to export data for this evaluation) which allows us to test the global effect of the AI-enabled self-referral tool on IAPT services. To date of this analysis (15/08/2022: note that NHS Digital UK only provides data with a delay so that this data set only included data up to May 2022), there were 4 IAPT providers with a total of 18 IAPT services that had implemented the AI-enabled self-referral tool and had used it for a sufficient amount of time in order to draw a pre-versus post -comparison (i.e. at least 4 months of data available post-implementation). The time of launch of the AI-enabled self-referral tool and the post-implementation period for the different IAPT providers can be seen in Figure 1A. For the timeframe in consideration, a total of 58,475 patients went through treatment at these IAPT services, and thus have been included in this analysis.

### 2.3 Definition of pre- and post -period

To conduct a pre-versus post-comparison for the AI-enabled self-referral tool, we chose a window of 3 months prior to the implementation of the tool to establish a reliable baseline for the recovery rate (see Figure 1A). We chose to use the month during which the tool was first adopted as part of this baseline period (e.g. if the tool has been launched on the 25th of July, treatment outcomes from July are still included in the pre-implementation baseline). This is because patients finishing their treatment during this first month will not have been referred through the self-referral tool and are thus representative of clinical assessments conducted prior to implementation of the tool.

The post-implementation period is intended to capture the effects of the AI-enabled self-referral tool on clinical assessments and subsequent clinical outcomes. Since the mean treatment time is 119 days from referral (data provided by one of the IAPT providers, based on treatment of *>* 20, 000 patients during the investigation period), patients are likely to only contribute to the treatment outcome measures around 4 months after they have been referred. Therefore, we should only see changes in treatment outcomes attributable to the AI-enabled self-referral tool 4 months after implementation. Thus, the post-period is defined from 4 months after implementation until the current date. The post-implementation periods included in the analysis for each IAPT provider were the following:

- IAPT provider 1 (7 IAPT services): 14 months (25,358 patients)
- IAPT provider 2 (2 IAPT services): 7 months (11,436 patients)
- IAPT provider 3 (2 IAPT services): 7 months (2,286 patients)
- IAPT provider 4 (7 IAPT services): 3 months (3,651 patients)

### 2.4 Outcome measure

The outcome measure of interest was reliable recovery [Jacobson and Truax, 1992]. This is routinely measured in IAPT by the relevant questionnaire measure of the patient’s mental health problem and is defined as a significant reduction in symptom scores from the beginning to the end of treatment, as well as a score below the clinical cut-off at the end of treatment. Importantly, the analysis shows similar results when a simple recovery rate was used as an outcome measure rather than a reliable recovery. This indicates that the conclusions are independent of the exact measure of recovery chosen. Both treatment outcome measures are reported in the NHS digital dataset.

### 2.5 Statistical Analysis

We compared recovery rates for IAPT services between the pre- versus post-implementation period for the IAPT services using the AI-enabled self-referral tool (see the four IAPT providers above). As such, a simple chi-square test was used to compare the frequency of recovered patients versus not recovered patients in the pre and post-implementation period. To control for the general effects of time and general pressure on the NHS we conducted the same analysis for all other NHS IAPT providers during the same time period.

### 2.6 Logistic regression analysis

In order to formally test a differential change in recovery rates from the pre- to the post-implementation period in IAPT providers using the AI-enabled self-referral tool versus other IAPT providers, we constructed a logistic regression predicting on a single patient level whether the patient recovered or not (0=not recovered, 1=recovered) as the dependent variable. This dependent variable was constructed based on the knowledge about how many people had recovered or not recovered for each provider each month. As predictor variables, we used time (0=pre-implementation, 1=post-implementation), whether the IAPT provider used the AI-enabled self-referral tool (0=no tool, 1=use of the AI-enabled self-referral tool) and their interaction. Hereby, we were specifically interested in the interaction effect as it captures a differential change in recovery rates from pre- to post-implementation for IAPT providers using the AI-enabled self-referral tool compared to the average IAPT. Finally, we also evaluated the recovery rates between the groups (0=no tool, 1=use of the AI-enabled self-referral tool) at the post-implementation time point to evaluate whether after the implementation of the AI-enabled self-referral tool the recovery rates were lifted above NHS average.

### 2.7 Control analysis

Since this was a quasi-experimental design, the IAPT services may have differed in some systematic ways which could affect our conclusions. Most importantly we wished to control for the recovery rate during the pre-implementation period as the baseline recovery could have a strong influence on the observed improvement, practically controlling for potential floor and ceiling effects. Moreover, IAPT services may have experienced changes in the number of treated patients (per month) from pre- to post-implementation period, which could affect the quality of care. Therefore, we wanted to ensure that these factors did not confound the observed effects.

We controlled for these potentially confounding effects in two ways: First by including them as covariates in the logistic regression and secondly by matching IAPT services using the AI tool to similar other IAPT services based on these characteristics.

The covariate analysis statistically accounts for linear effects of baseline recovery rate and changes in treated patients, but not non-linear effects. Therefore, in the second step of control analyses, we additionally matched the IAPT services using the AI tool with other IAPT services (not using the AI tool) which were similar in terms of these characteristics. This matching is similar to a matched-control analysis commonly employed in clinical and public health studies [Rose and Van der Laan, 2009]. For each IAPT service using the AI tool, we calculated recovery rates during the pre-implementation period and changes in the number of treated patients from pre- to post-implementation period. We calculated the same metrics for all other IAPT services during the same period of time. In order to match services, we calculated the Euclidean distance based on these two (normalized) features and took the services with the lowest distance to IAPT services using the AI tool, where a low distance implies a high degree of similarity. Given these matched comparison groups we then conducted the logistic regression as outlined above.

In the first analysis, we chose for each IAPT provider using the AI tool one most similar IAPT provider (not using the AI tool) as a comparison. To ensure that these results were not conflated by the specific comparison group chosen, we also compared the IAPT services using the AI tool to the 25% and 50% of IAPT services which were most similar to them (based on the Euclidean distance described above) ensuring that the results are not dependent on the exact comparison group.

#### 3 Results

### 3.1 Recovery rates

A total of 58,360 patients were referred via the AI-enabled self-referral tool (*Limbic Access*) to IAPT services during the timeframes under investigation. This included 15,629 in the pre-implementation period and 42,731 in the post-implementation period (see Methods). The recovery rate was lower in the pre-implementation period (47.1%) compared to the post-implementation period (48.9%). The increased recovery rate in the post-implementation group represented a significant improvement (Odds-ratio=1.078, CI=[1.039, 1.119], *χ*=16.15, *p <* .00001). Figure 1C shows the change in recovery rates over time relative to the implementation of the AI-enabled self-referral tool.

It must be noted that the nature of a pre- versus post-implementation study introduces the possibility of confounding factors that could could explain the observed effects (e.g. other variables could have changed during the investigated timeframe). For instance, general pressure on IAPT services could have eased, or other factors could have caused a more beneficial outcome of treatment during the post-implementation period.

Importantly, the NHS Digital dataset provides an avenue to control for confounding influences as we can investigate the treatment outcomes reported by other IAPT services (which did not implement an AI self-referral tool) during the same time. This allows us to control for general changes regarding pressure on IAPT services or any other effect that is driven by the difference in time.

Investigating the same timeframe in all other IAPT services (those not implementing the AI solution) showed that overall 889,934 patients were treated in these services: 148,790 in the pre-implementation period and 741,144 in the post-implementation period. In these IAPT services, the recovery rate decreased from the pre- (48.3%) to the post-implementation period (46.9%) (see Figure 2), which represents a highly significant reduction in recovery rate (Odds-ratio=0.945, CI=[0.936, 0.957], *χ*=94.5, *p <* .00001).

**Figure 2:**
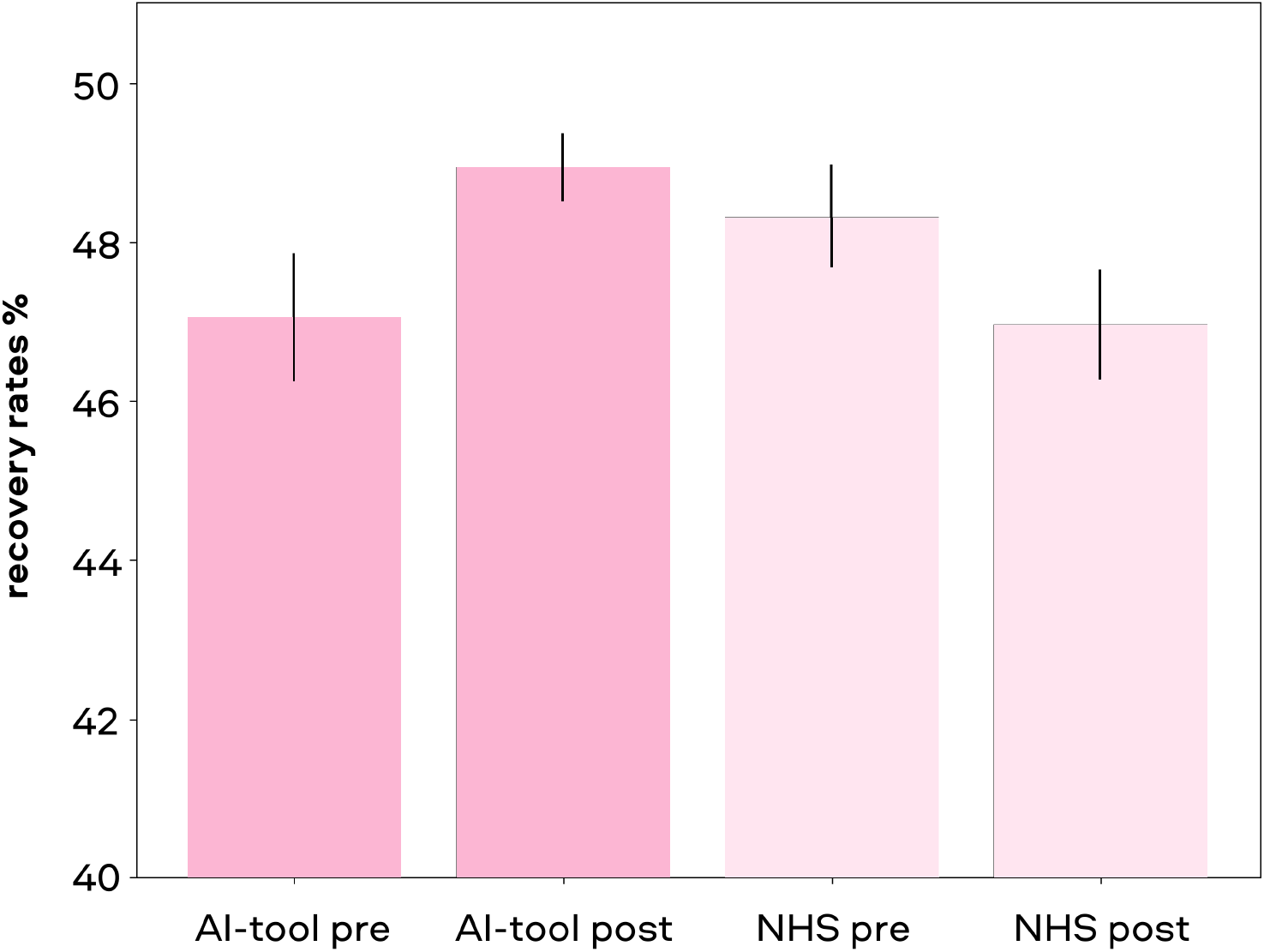
Comparison of recovery rates in the pre versus post AI tool implementation period for IAPT services that used the referral tool compared to the NHS average. Error bars indicate standard errors between IAPT services in each group.

While the IAPT services which used the AI-enabled self-referral tool had improved recovery rates during the time of investigation, other IAPT services showed a reduction in recovery rates. This indicates that the improvement seen with the AI tool cannot be explained by temporal effects alone (e.g. data acquired during the COVID-19 pandemic), as the effect would then also be apparent in the other IAPT services. A formal test of this differential effect of time on recovery rates is an interaction effect between time and the presence of the AI-enabled self-referral tool. As expected, we found a highly significant interaction term (*β*=.219, z=11.083, *p <* .00001), indicating that IAPT services using the AI tool showed significantly higher improvements in recovery from the pre- to post-implementation period. Importantly, when only focusing on the post-implementation phase, IAPT services using the AI tool showed significantly better recovery rates than IAPT services not using the tool (*β*=.06, z=6.36, *p <* .00001). This indicates that the usage of the AI-enabled self-referral tool boosted the quality of care above the NHS average.

However, there might have been some more general differences between the average NHS IAPT service and the services using the AI tool. For instance, it is apparent that the recovery rate during the pre-implementation period was different between IAPTs that use the AI-enabled self-referral tool and the average NHS IAPT, which could for instance lead to regression to the mean effects. Moreover, the two groups may have differed in the number of patients they treated and how this changed from pre- to post-implementation period which could have affected the changes in recovery rates. For this reason, we included the recovery at pre-implementation as well as change in treated patients (from pre to post) as covariates in our analysis reported above to statistically account for these potentially confounding variables. Importantly, these effects did not change the interaction effect and could thus not explain the observed effects of the AI-enabled self-referral tool (*p <* .00001).

While statistically controlling for these differences is the first step, it is possible to better control for these potential confounds by only comparing the IAPT services which used the AI tool against other IAPT services that are closely matched for recovery rates at pre-implementation baseline and changes in the number of patients treated from pre- to post-period. Therefore, in addition to statistically controlling for potentially confounding effects, we also matched our NHS control group based on these covariates.

First, we chose the single most similar IAPT service for each of the AI IAPTs. This led to comparable initial recovery rates for the IAPTs using the (47.1%) and the comparison group (47.6%). Similarly, the groups were well matched regarding their change in treated patients, whilst AI IAPTs showed a reduction in treated patients from pre- to post-implementation of 2.1%, the comparison group showed a change of 1.8%. This indicates that the matching of comparison groups worked well. Importantly, conducting the same analysis as before, we still found a significant interaction effect (*β*=.14, z=3.69, *p <* .00001) meaning that IAPTs using the AI tool improved their recovery rates more than the comparison group (see Figure 3A).

**Figure 3:**
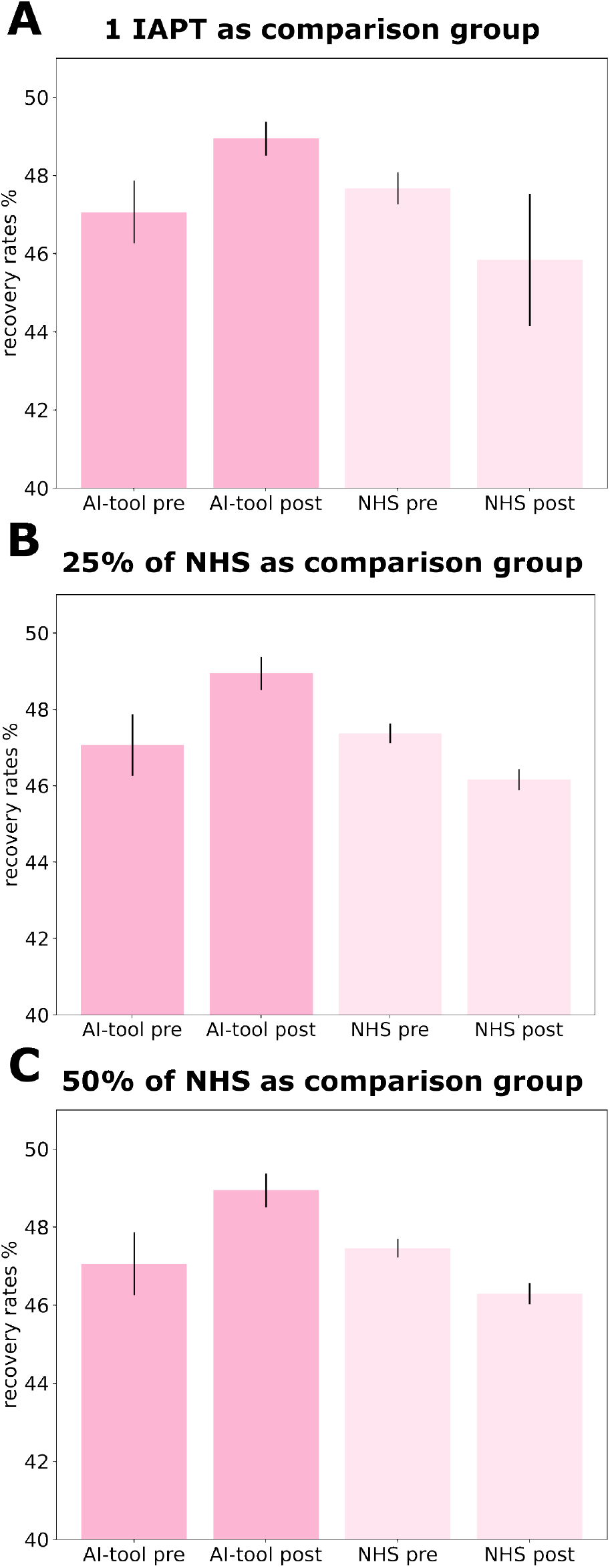
Comparison of recovery rates in the pre versus post AI implementation period for IAPT services that used the AI-enabled self-referral tool compared to the NHS IAPT services with matched characteristics. A) Comparison against the most similar IAPT without AI. B) Comparison against the 25% (of all NHS IAPTs) most similar IAPTs without AI. B) Comparison against the 50% (of all NHS IAPTs) most similar IAPTs without AI. Error bars indicate standard erros between IAPT services in each group.

Nevertheless, comparing the IAPT services using the AI tool only against the most similar IAPT service might introduce some sampling bias as this analysis is highly dependent on the chosen comparison group. Therefore, we decided to compare those against a larger proportion of the NHS IAPT services while still matching for initial recovery and changes in the number of treated patients. When comparing each IAPT service to either the 25% most similar or the 50% most similar NHS IAPT services in terms of recovery and change of treated patients, we still found significant interaction effects between AI tool usage and time (25% IAPT services: *β*=.125, z=6.38, *p <* .00001; 50% IAPT services: *β*=.158, z=7.94, *p <* .00001) with a stronger improvement in recovery rate for IAPTs using the AI tool than in the comparison group (see Figures 3B and 3C).

#### 3.2 Economic analysis

#### 3.2.1 Cost per recovery associated with AI triage

Beyond clinical efficacy, AI solutions have the potential yield economic benefit for healthcare delivery. In an effort to evaluate the cost-effectiveness of AI-enabled triage and assessment support chatbots in psychological therapy, we evaluated the associated cost for each additional recovery associated with the AI solution (*Limbic Access*) in IAPT.

As shown in 3C, utilisation of the AI-enabled self-referral tool was associated with an improvement in recovery rates of 3.1% compared to the matched NHS services in the same time period. Taking the number of patients treated in the IAPT services using the AI tool during the post-implementation period (42,731), combined with the associated 3.1% improvement in recovery rate, it is reasonable to estimate 1,304 additional recovered patients in these IAPT services associated with the usage of the AI-enabled triage and assessment support. Price per recovery can then be estimated by dividing the cost of implementing the AI solution by the number of additionally recovered patients.

It is important to note that the referrals to IAPT services using the AI tool are not made exclusively via the AI-enabled self-referral tool itself. Indeed, to promote patients choice and mitigate digital exclusion, IAPT services are encouraged to maintain alternative avenues for referral (e.g. telephone referrals). During the time of analysis, a total of 27,029 patient referrals into IAPT services were processed via the AI tool. The commercial costs associated with implementing and running the AI tool in IAPT services during the evaluation period is outlined in Table1. A tiered pricing structure indicates a combined cost to IAPT services between £135,145 and £270,290 to process 27,029 patient referrals via the AI tool. Dividing this cost by the number of additionally recovered patients yields the cost associated with each additional recovery: The lower bound for the price per referral (£5 per processed referral) results in a cost of £103.64 per additional recovery (£135,145/1,304). In comparison, a higher price per processed referral (£10 per processed referral) would result in an upper bound of £207.28 per additional recovery (£270,290/1,304) for the AI-enabled self-referral tool.

**Table 1:** Price per additional recovery based on the tiered pricing structure for the AI-enabled self-referral tool

| Price per referral (£) | Price overall (£) | Price per additional recovery (£) |
| --- | --- | --- |
| 5 | 135,145 | 103.64 |
| 7.5 | 202,718 | 155.46 |
| 10 | 270,290 | 207.28 |

#### 3.2.2 A brief comparison with alternative options to improve recovery

It is important to compare the associated cost per recovery of the AI-enabled self-referral tool (outlined in Table1) with alternative approaches to improve recovery rates. One option would be to invest in more mental health practitioners to provide additional therapy sessions for every patient in treatment. It has been shown that every additional session of low-intensity therapy within the IAPT care system increases the likelihood of recovery with an odds ratio of 1.204 Gyani et al. [2013]. Taking the existing baseline recovery rate in IAPT, an additional treatment session for every patient would therefore be expected to improve overall recovery rates by 5.1%. Applying this to the 27,029 patients in our dataset, a 5.1% improvement in recovery rates would yield 2,711 additionally recovered patients. However, each session of therapy is very costly - one low-intensity therapy session in IAPT has been estimated to cost £102.38 [Griffiths and Steen, 2013]. Offering each of the 27,029 patients in our dataset one additional therapy session would therefore amount to a total cost of £2,767,229.02 (1,021 per additional recovery). This is 5x higher than even the upper bound on cost per recovery for the AI-enabled self-referral tool. Furthermore, existing recruitment limitations casts doubt on the feasibility of reliably increasing the clinical workforce to deliver additional treatment sessions for every patient in IAPT.

Another option would be to use alternative digital tools to support clinical assessments. To our knowledge, only one study conducted a clinical efficiency analysis for digital tools supporting mental health assessment in IAPT. In this study, Delgadillo et al. [2022] developed a machine-learning algorithm which was used to support the clinical assessment in IAPT. This model predicted which treatment intensity would likely result in recovery for a given patient (IAPT operates on a stepped care system). Usage of this tool improved recovery rates, however it also resulted in an increased number of patients being treated in a higher level of treatment intensity (creating some additional cost).

The authors also conducted an economic analysis, estimating a cost of £1,320 per additional recovery for their solution. Again, this is substantially higher than the cost per recovery for the AI-enabled self-referral tool evaluated in this study.

Taken together, this analysis provides strong evidence to suggest that the investigated AI-enabled self-referral tool represents an economically viable avenue to improve recovery rates in psychological therapy, as indicated in the NHS IAPT care system.

## 4 Discussion

In summary, we found that IAPT providers using an AI-enabled self-referral tool to conduct self-referrals Limbic Access) showed a significant increase in recovery rates after the launch of this tool. In contrast, comparable NHS IAPT services that did not use this tool showed a decline in recovery rates during the same time period. We conducted multiple control analyses to ensure that this effect cannot be explained by potentially confounding factors, such as general time effects or differences in IAPT services. Across all analysis, our results clearly show that this AI-enabled self-referral tool has a positive effect on patient recovery rates.

One caveat that should be considered is a possible difference in culture between different IAPT services, which is not reflected in patient recovery at baseline or admission rates. For example, the IAPTs using the AI tool may inherently be more open to digital health approaches and innovation compared to IAPTs not adopting such tools, which in turn may affect recovery rates. We believe, however, that this is an unlikely scenario because our analyses investigated a change in recovery that coincided with the launch of the AI tool (cf Figure 1A). Any cultural differences between IAPT services would have an effect on the general recovery rate, and not to the change of those time-locked to the launch of the AI tool. We thus deem this alternative as highly unlikely.

Because we used a pre- versus the post-implementation period design and compare the changes to matched IAPT services, we can be confident that the observed are not due to general effects, such as external pressure on IAPT services or any other general and time-dependent factors.

Here, we used a large dataset provided for all IAPT services across the UK. This allowed us to assess the effects on a large-scale level and thus reduce common biases that often occur in smaller sample studies. However, given that these are not patient specific data, it is not possible to investigate more fine-grained mechanisms of how and why exactly the introduction of a conversational AI tool improves recovery rates. This should be investigated in more targeted future studies. We cannot clearly disentangle whether these improved recovery rates are a direct consequence of the AI tool improving the quality of the clinical assessment or whether these observed improvements are indirect effects of driving clinical efficiencies and through this freeing up the time of clinicians which then results in improved quality of treatment and increased recovery rates. Given that there are many intermediate steps that influence treatment outcomes, it is remarkable that we could reliably find highly significant effects on recovery rates. However, other internal evaluations on this AI-enabled self-referral tool have indicated that the effect is driven by a direct effect through an improvement of the assessment process (in prep).

Lastly, we calculated the cost-effectiveness of the AI-enabled self-referral tool and estimated that to be £103.64 - £207.3 per additional recovery (depending on the exact price point) which represents an extremely cost-effective way of improving recovery rates, being an order of magnitude more cost-effective than the next best solution. Interestingly, the cost savings to the NHS for each additional person recovered from mental health problems have been estimated to be £300 over a period of 2 years whereby the overall cost to the economy has been estimated to be £1200 per person within 2 years [Laynard et al., 2007]. This indicates that AI tools might provide a highly positive cost-benefit ratio for the NHS and society as a whole.

In summary, these results indicate that the usage of AI-enabled referral tools create clinical efficacy and improve treatment outcomes for patients. Moreover, it appears that this solution represents a cost-effective way of achieving these improvements, further suggesting a large potential for the adoption of this technology.

## Data Availability

Code and data supporting this study are available at a dedicated Github repository (https://github.com/LimbicAI/limbic-NHSDigital-RecoveryRates.git)

https://github.com/LimbicAI/limbic-NHSDigital-RecoveryRates

## 5 Acknowledgments

We thank Emili Ivanova for her contribution to the figures and illustrations in this research article.

## 7 Data and code availability

Code and data supporting this study are available at a dedicated Github repository.

## References

R. Adams, T. Ryan, and E. Wood. Understanding the factors that affect retention within the mental health nursing workforce: a systematic review and thematic synthesis. International Journal of Mental Health Nursing, 30(6):1476–1497, 2021.

G. Busetta, M. G. Campolo, F. Fiorillo, L. Pagani, D. Panarello, and V. Augello. Effects of covid-19 lockdown on university students’ anxiety disorder in italy. Genus, 77(1):1–16, 2021.

L. T. Car, D. A. Dhinagaran, B. M. Kyaw, T. Kowatsch, S. Joty, Y.-L. Theng, R. Atun, et al. Conversational agents in health care: scoping review and conceptual analysis. Journal of medical Internet research, 22(8):e17158, 2020.

K. Ćsić, S. Popović, M. Šarlija, and I. Kesed žić. Impact of human disasters and covid-19 pandemic on mental health: potential of digital psychiatry. Psychiatria Danubina, 32(1):25–31, 2020.

J. Delgadillo, S. Ali, K. Fleck, C. Agnew, A. Southgate, L. Parkhouse, Z. D. Cohen, R. J. DeRubeis, and M. Barkham. Stratified care vs stepped care for depression: A cluster randomized clinical trial. JAMA psychiatry, 79(2):101–108, 2022.

S. D’Alfonso. Ai in mental health. Current Opinion in Psychology, 36:112–117, 2020.

K. K. Fitzpatrick, A. Darcy, and M. Vierhile. Delivering cognitive behavior therapy to young adults with symptoms of depression and anxiety using a fully automated conversational agent (woebot): A randomized controlled trial. JMIR mental health, 4, 2017. doi: 10.2196/MENTAL.7785. URL https://pubmed.ncbi.nlm.nih.gov/28588005/.

S. Griffiths and S. Steen. Improving access to psychological therapies (iapt) programme: Scrutinising iapt cost estimates to support effective commissioning. The Journal of Psychological Therapies in Primary Care, 2(2):142–156, 2013.

A. Gyani, R. Shafran, R. Layard, and D. M. Clark. Enhancing recovery rates: lessons from year one of iapt. Behaviour research and therapy, 51(9):597–606, 2013.

T. U. Hauser, V. Skvortsova, M. De Choudhury, and N. Koutsouleris. The promise of a model-based psychiatry: building computational models of mental ill health. The Lancet Digital Health, 2022.

N. S. Jacobson and P. Truax. Clinical significance: a statistical approach to defining meaningful change in psychotherapy research. 1992.

K. Jayaraajan, A. Sivananthan, A. Koomson, A. Ahmad, M. Haque, and M. Hussein. The use of digital solutions in alleviating the burden of iapt’s waiting times. International Journal of Risk & Safety in Medicine, (Preprint):S1–S8, 2022.

N. Koutsouleris, T. U. Hauser, V. Skvortsova, and M. De Choudhury. From promise to practice: towards the realisation of ai-informed mental health care. The Lancet Digital Health, 2022.

K. Kroenke, R. L. Spitzer, and J. B. Williams. The phq-9: validity of a brief depression severity measure. Journal of general internal medicine, 16(9):606–613, 2001.

P. Larsson, R. Lloyd, E. Taberham, and M. Rosairo. An observational study on iapt waiting times before, during and after the covid-19 pandemic using descriptive time-series data. Mental Health Review Journal, (ahead-of-print), 2022.

R. Laynard, D. Clark, M. Knapp, and G. Mayraz. Cost-benefit analysis of psychological therapy. National Institute Economic Review, 202(1):90–98, 2007.

A. M. Loosen, V. Skvortsova, and T. U. Hauser. Obsessive–compulsive symptoms and information seeking during the covid-19 pandemic. Translational psychiatry, 11(1):1–10, 2021.

L. Marques, A. D. Bartuska, J. N. Cohen, and S. J. Youn. Three steps to flatten the mental health need curve amid the covid-19 pandemic. Depression and Anxiety, 37(5):405, 2020.

R. Meadows, C. Hine, and E. Suddaby. Conversational agents and the making of mental health recovery. Digital Health, 6, 2020. doi: 10.1177/2055207620966170. URL /pmc/articles/PMC7683843//pmc/articles/PMC7683843/?report=abstracthttps://www.ncbi.nlm.nih.gov/pmc/articles/PMC7683843/.

B. Murch, J. Cooper, T. Hodgett, E. Gara, J. Walker, and R. Wood. Modelling the effect of first-wave covid-19 on mental health services. Operations Research for Health Care, 30:100311, 2021.

S. Nochaiwong, C. Ruengorn, K. Thavorn, B. Hutton, R. Awiphan, C. Phosuya, Y. Ruanta, N. Wongpakaran, and T. Wongpakaran. Global prevalence of mental health issues among the general population during the coronavirus disease-2019 pandemic: a systematic review and meta-analysis. Scientific Reports, 11(1):1–18, 2021.

F. Ornell, W. V. Borelli, D. Benzano, J. B. Schuch, H. F. Moura, A. O. Sordi, F. H. P. Kessler, J. N. Scherer, and L. von Diemen. The next pandemic: impact of covid-19 in mental healthcare assistance in a nationwide epidemiological study. The Lancet Regional Health-Americas, 4:100061, 2021.

S. Rose and M. J. Van der Laan. Why match? investigating matched case-control study designs with causal effect estimation. The international journal of biostatistics, 5(1), 2009.

B. N. Rudd and R. S. Beidas. Digital mental health: the answer to the global mental health crisis? JMIR Mental Health, 7(6):e18472, 2020.

M. Scott. The cost of iapt is at least five times greater than claimed. CBT watch, 2018a.

M. J. Scott. Improving access to psychological therapies (iapt)-the need for radical reform. Journal of health psychology, 23(9):1136–1147, 2018b.

R. L. Spitzer, K. Kroenke, J. B. Williams, and B. Löwe. A brief measure for assessing generalized anxiety disorder: the gad-7. Archives of internal medicine, 166(10):1092–1097, 2006.

J. Thome, J. Deloyer, A. N. Coogan, D. Bailey-Rodriguez, O. A. da Cruz E Silva, F. Faltraco, C. Grima, S. O. Gudjonsson, C. Hanon, M. Holly’, et al. The impact of the early phase of the covid-19 pandemic on mental-health services in europe. The World Journal of Biological Psychiatry, 22(7):516–525, 2021.

A. N. Vaidyam, H. Wisniewski, J. D. Halamka, M. S. Kashavan, and J. B. Torous. Chatbots and conversational agents in mental health: A review of the psychiatric landscape. The Canadian Journal of Psychiatry, 64:456–464, 2019. doi: 10.1177/0706743719828977.

L. Wilson, M. Marasoiu, et al. The development and use of chatbots in public health: Scoping review.JMIR human factors, 9(4):e35882, 2022.

